# Hepatitis B surface antigen clearance in HIV/HBV-coinfected patients: associated clinical factors and a rare amino acid substitution underlying occult infection

**DOI:** 10.64898/2026.09.02.26362028

**Authors:** Manabu Takabayashi, Tomofumi Nakamura, Hirotomo Nakata, Jun-ichirou Yasunaga, Shuzo Matsushita

## Abstract

**Background:** HBV coinfection is more common among people with HIV than in the general population. Nucleotide reverse transcriptase inhibitors (NRTIs) in antiretroviral therapy (ART) are also active against HBV. Growing use of NRTI-sparing regimens raises concern about HBV reactivation.

**Objectives:** To determine the rate, cumulative incidence, and factors associated with HBsAg clearance in HIV/HBV-coinfected patients, and to characterize the genetic/structural basis of occult HBV infection (OBI) in the same cohort.

**Study design:** We retrospectively reviewed 256 people with HIV at Kumamoto University Hospital in Japan (1986–2025), stratified by HBsAg clearance status; clinical factors were compared and cumulative incidence estimated by Kaplan–Meier analysis. OBI cases underwent HBsAg-region sequencing.

**Results:** Fifteen patients were analyzed (12 HBsAg-positive, 3 OBI). HBsAg clearance occurred in 7/12 (58%). The Kaplan–Meier estimated cumulative incidence reached 25% by 2 years and 64.3% at approximately 4.4 years, after which no additional clearance events were observed during follow-up. Clearance was associated with higher CD4 (≥ 200 cells/μL: 71% vs 0%) and lower HIV-RNA (<10^5^ copies/mL: 86% vs 20%); HBV-DNA was significantly higher in the clearance group than OBI (adjusted *p* = 0.007). All OBI sequences matched genotype C2; one also carried a rare genotype A2 ‘a’ determinant substitution, N131K (0.23% of genotype A sequences in HBVdb).

**Conclusions:** HBsAg clearance was frequent, concentrated within 3–4 years of ART, and associated with preserved immune status and HBV replicative activity. A rare ‘a’ determinant substitution may cause false-negative HBsAg in OBI, informing NRTI-sparing ART selection.

**Highlights:**

- No further HBsAg clearance events were observed after approximately 4.4 years.
- Higher CD4 count and lower HIV-RNA at baseline were associated with HBsAg clearance.
- HBV-DNA was significantly higher in HBsAg-clearance patients than in OBI patients.
- A rare ‘a’ determinant substitution (N131K) was found in one occult infection case.
- *In silico* modeling suggested that N131K may perturb HBsAg–antibody binding affinity through altered loop dynamics, not direct epitope disruption.

## Background

The prevalence of hepatitis B virus (HBV) infection is substantially higher among people living with HIV than in the general population; in Japan, HBsAg positivity has been reported at 6.4% among HIV-infected individuals overall and 8.3% among men who have sex with men, compared with 0.28% in the general population [1,2]. This elevated prevalence reflects shared transmission routes and is clinically consequential because standard antiretroviral therapy (ART) has historically included two nucleotide reverse transcriptase inhibitors (NRTIs), most commonly tenofovir or lamivudine, both of which are dually active against HIV and HBV. Under NRTI-containing ART, HIV treatment has therefore functioned simultaneously as de facto HBV treatment.

Advances in antiretroviral drug design have shifted this paradigm. Two-drug regimens that reduce or eliminate NRTIs (such as dolutegravir plus lamivudine, or dolutegravir plus rilpivirine) are now widely used to limit long-term NRTI-related toxicity (nephrotoxicity, reduced bone mineral density). However, removing NRTI-based HBV suppression has unmasked HBV reactivation in patients previously controlled incidentally by HIV treatment [3–7]. Stratifying reactivation risk before such regimen switches requires assessing HBV serological markers, but the appropriate approach for patients with isolated anti-HBc positivity, or for those with occult HBV infection (OBI; defined as HBsAg-negative but HBV-DNA-detectable) remains unclear, partly because OBI itself is difficult to diagnose in patients already receiving NRTI-containing ART, limiting epidemiological data [1,8,9,10].

Our institution is a designated AIDS treatment core hospital in Japan and follows a large single-center HIV cohort, offering an opportunity to characterize both the prevalence of OBI and the clinical course of HBsAg-positive coinfection, including the rate and determinants of spontaneous or treatment-associated HBsAg clearance, information directly relevant to decisions about NRTI-sparing ART.

### Objectives

This study aimed to (i) determine the rate and cumulative incidence of HBsAg clearance among HIV/HBV-coinfected patients and identify associated clinical factors, and (ii) characterize the HBV genetic and structural basis of OBI cases identified within the same cohort, to help define which HBV serological profiles may safely permit NRTI-sparing ART.

### Study design

This retrospective observational study was approved by the institutional review board of Kumamoto University Hospital (approval no. 1942). We reviewed all people living with HIV followed at our hospital as of January 2025 (n=256; cohort accrued 1986–2025). HIV/HBV coinfection was defined as HBsAg positivity or detectable HBV-DNA at the time of HIV diagnosis, with OBI defined as HBsAg-negative status with detectable HBV-DNA. Patients with insufficient clinical data, acute HBV infection, or HBV-specific antiviral treatment predating HIV diagnosis were excluded. The observation period extended from the first clinic visit to the most recent blood test.

Among HBsAg-positive patients, HBsAg clearance was defined as seroconversion to HBsAg-negative status confirmed on follow-up testing. Clearance and non-clearance groups were compared for CD4 count, HIV-RNA level, HBV-DNA level, ALT, and HBeAg status at baseline. These continuous variables, including age, BMI, CD4 count, HIV-RNA level, HBV-DNA level, and ALT, were compared across the three groups using the Kruskal–Wallis rank-sum test. Pairwise comparisons were performed using Dunn’s test with Bonferroni adjustment. Categorical variables were compared using Fisher’s exact test [11]. All tests were two-sided, and a Bonferroni-adjusted p value <0.05 was considered statistically significant; given the small sample size, analyses were considered exploratory. Cumulative incidence of HBsAg clearance was estimated using the Kaplan–Meier method [12] with time zero at ART initiation. For OBI cases, residual serum obtained at first presentation was used to amplify the HBsAg-coding region (approximately 1.2 kb within the ∼3.2-kb HBV genome) by PCR, followed by Sanger sequencing. Deduced amino acid sequences were aligned against reference genotype C2 (GenBank AB050018) and genotype A2 (GenBank AB116080) sequences, with particular attention to the major hydrophilic region (MHR) and, within it, the ‘a’ determinant (residues 124–146) of HBsAg, the principal epitope recognized by most commercial HBsAg assays. Residue frequency at variant positions was cross-referenced against the HBVdb database [13].

For *in silico* structural analysis of residue 131, the cryo-EM structure of HBsAg in complex with the HBC34 neutralizing antibody Fab fragment (PDB 9U9B) was used as the structural template. HBsAg forms a homodimer; inspection of all three available Fab-complex structures (9U9B, 9IYY, 9JT1) revealed complementary electron density gaps in the two monomers; residues approximately 127–132 were unresolved in one monomer, while residues approximately 110–116 were unresolved in the partner monomer in which residue 131 was structurally defined. The latter monomer was selected for analysis. The unresolved 110–116 loop was completed by MODELLER loop refinement (10 candidate models; best selected by internal energy score) [19]. *In silico* point mutations to asparagine (N131) and lysine (K131) were introduced using FoldX (BuildModel, five independent runs each). Molecular dynamics (MD) simulations were performed using GROMACS 2021.4 [20] with the AMBER ff14SB force field and TIP3P water model. Each system (∼76,000 atoms) was solvated in a dodecahedral box with 150 mM NaCl, energy-minimized by steepest descent, equilibrated under NVT (100 ps) and NPT (100 ps) conditions, and subjected to 50 ns production simulation. Binding free energies (MM-GBSA, without entropy correction) were calculated from the last 10 ns of each simulation (101 equally spaced frames at 0.1 ns intervals) using gmx_MMPBSA [21,22]. Statistical comparison of ΔG_bind_ across constructs was performed using Welch’s t-test on independent simulation replicates.

## Results

Of 256 people living with HIV, 19 had HIV/HBV coinfection; after excluding 1 patient with insufficient data, 1 with acute HBV infection, and 2 with HBV treatment predating HIV diagnosis, 15 patients were analyzed: 12 HBsAg-positive and 3 with OBI (**Figure 1**). All 15 patients were male; mean age was 38±11 years. The prevalence of OBI in the HIV-positive cohort was 3 of 256 people with HIV (1%; 95% CI 0.3–3.1%).

**Figure 1.**
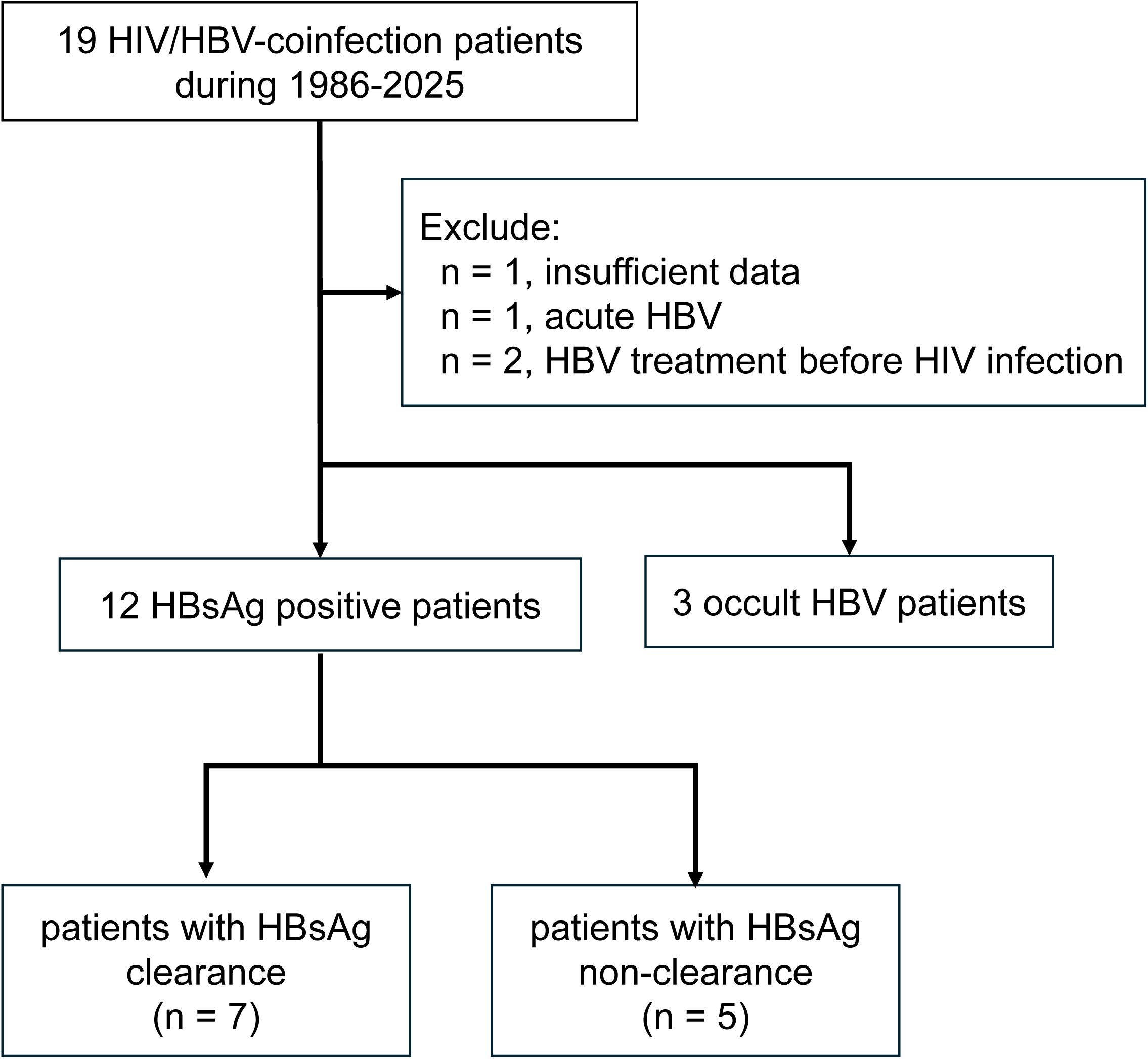
Patient flow diagram. Of 19 HIV/HBV-coinfected patients identified between 1986 and 2025, 4 were excluded (insufficient data, n = 1; acute HBV, n = 1; HBV treatment before HIV diagnosis, n = 2), leaving 15 patients for analysis: 12 HBsAg-positive (7 with HBsAg clearance, 5 with HBsAg non-clearance) and 3 with occult HBV infection (OBI).

Table 1 summarizes baseline characteristics of the cohort, stratified by HBsAg clearance status and OBI. Patients with OBI were, on average, older (43 ± 15 years) than those with HBsAg clearance (36 ± 11 years) or HBsAg non-clearance (39 ± 8 years), and had a higher body mass index than the clearance group (23.1 ± 2.2 vs 19.8 ± 3.1 kg/m^2^).

**Table 1.** Characteristics of HBsAg clearance and OBI in PLWH.

| Characteristic | Overall<br>(n = 15) | HBsAg clearance<br>(n = 7) | HBsAg non-clearance<br>(n = 5) | OBI<br>(n = 3) |
| --- | --- | --- | --- | --- |
| Age, years, mean (SD) | 38 (11) | 36 (11) | 39 (8) | 43 (15) |
| Male sex | 15 (100%) | 7 (100%) | 5 (100%) | 3 (100%) |
| BMI, kg/m<sup>2</sup>, mean (SD) | 21.6 (3.9) | 19.8 (3.1) | 23.6 (4.6) | 23.1 (2.2) |
| HIV-RNA <10<sup>4</sup> copies/mL | 7 (47%) | 6 (86%) | 1 (20%) | 0 (0%) |
| HIV-RNA ≥10<sup>4</sup> copies/mL | 8 (53%) | 1 (14%) | 4 (80%) | 3 (100%) |
| CD4 <200 cells/μL | 8 (53%) | 2 (29%) | 5 (100%) | 1 (33%) |
| CD4 ≥200 cells/μL | 7 (47%) | 5 (71%) | 0 (0%) | 2 (67%) |
| ALT, U/L, median (Q1–Q3) | 30 (24-63) | 44(23-94) | 30(28-34) | 28(19-30) |
| HBV-DNA <5 log IU/mL | 6 (40%) | 1 (14%) | 2 (40%) | 3 (100%) |
| HBV-DNA ≥5 log IU/mL | 9 (60%) | 6 (86%) | 3 (60%) | 0 (0%) |
| HBeAg negative | 3 (20%) | 1 (14%) | 1 (20%) | 1 (33%) |
| HBeAg positive | 10 (67%) | 6 (86%) | 4 (80%) | 0 (0%) |
| HBeAg unknown | 2 (13%) | 0 (0%) | 0 (0%) | 2 (67%) |

PLWH, people with HIV; OBI, occult HBV infection; BMI, body mass index; ALT, alanine aminotransferase; Q1–Q3, first and third quartiles.

Among the 12 HBsAg-positive patients, HBsAg clearance occurred in 7 (58%; 95% CI 32.0–80.7%) during follow-up. The Kaplan–Meier estimated cumulative incidence increased from ART initiation, reaching 25% by 2 years and 64.3% at approximately 4.4 years. No additional clearance events were observed thereafter during up to 14 years of follow-up (**Figure 2**), suggesting that the window for HBsAg clearance is established early after treatment initiation.

**Figure 2.**
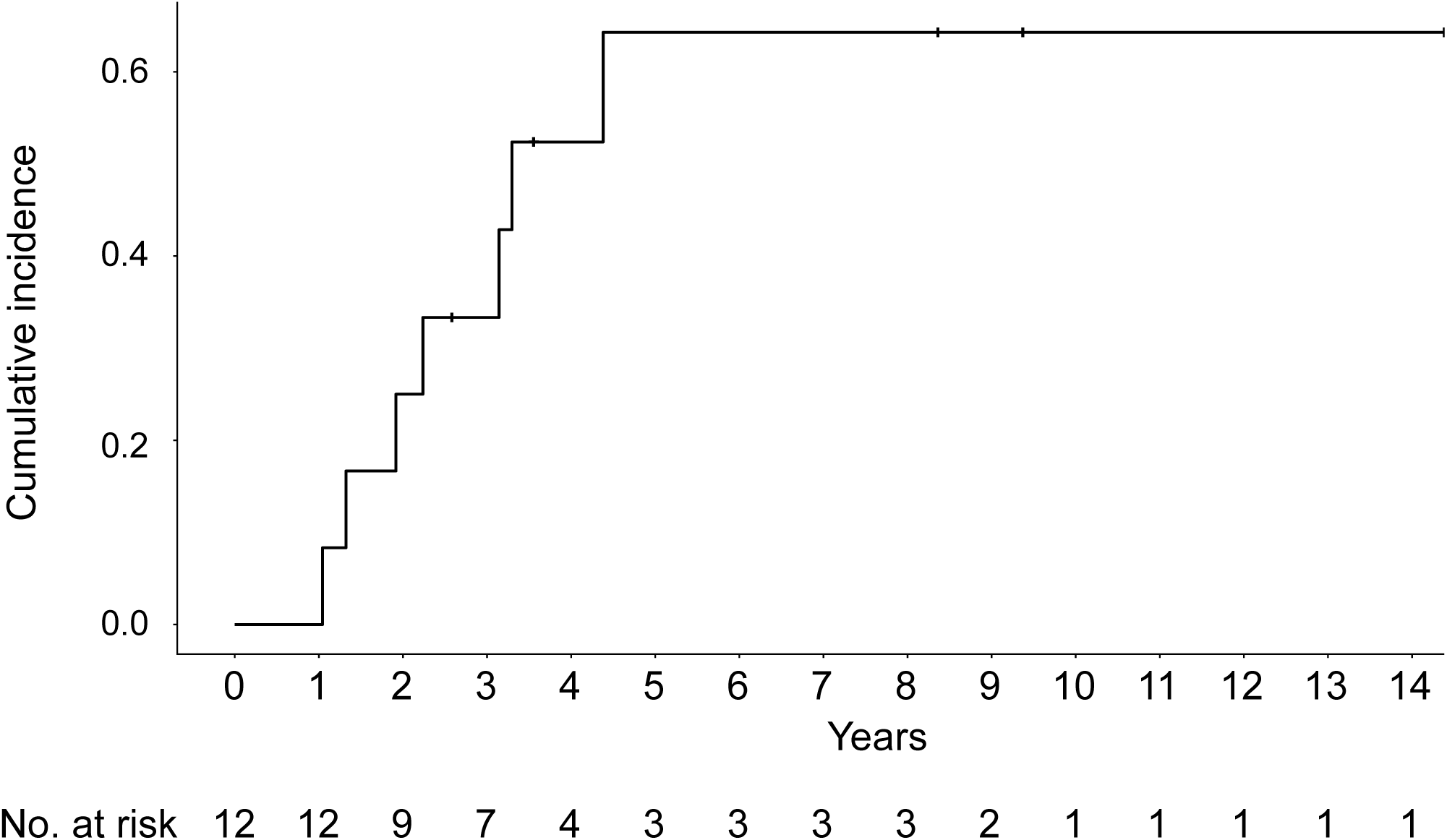
Cumulative incidence of HBsAg clearance among 12 HBsAg-positive HIV-coinfected patients (Kaplan–Meier estimate), with number at risk shown below the time axis.

Patients who cleared HBsAg more often had CD4 ≥ 200 cells/μL at baseline than those in the non-clearance group (71% vs 0%) and were more often HIV-RNA <10^5^ copies/mL (86% vs 20%) (**Table 1**). Comparing HBV-DNA, ALT, HIV-RNA, and CD4 across the three groups (HBsAg clearance, HBsAg non-clearance, and OBI), HBV-DNA differed significantly between the HBsAg clearance and OBI groups (median markedly higher in the clearance group; *p* = 0.007), while the remaining pairwise HBV-DNA comparisons, and all comparisons for ALT, HIV-RNA, and CD4, did not reach significance (**Figure 3**), likely reflecting the small sample size. Despite the lack of statistical significance, CD4 counts were much higher in the clearance group (median ∼310 cells/μL) than in the non-clearance group (∼75 cells/μL). HBeAg was positive in 10/15 patients overall (67%; 86% of the clearance group and 80% of the non-clearance group) and could not be determined in 2 of the 3 OBI cases.

**Figure 3.**
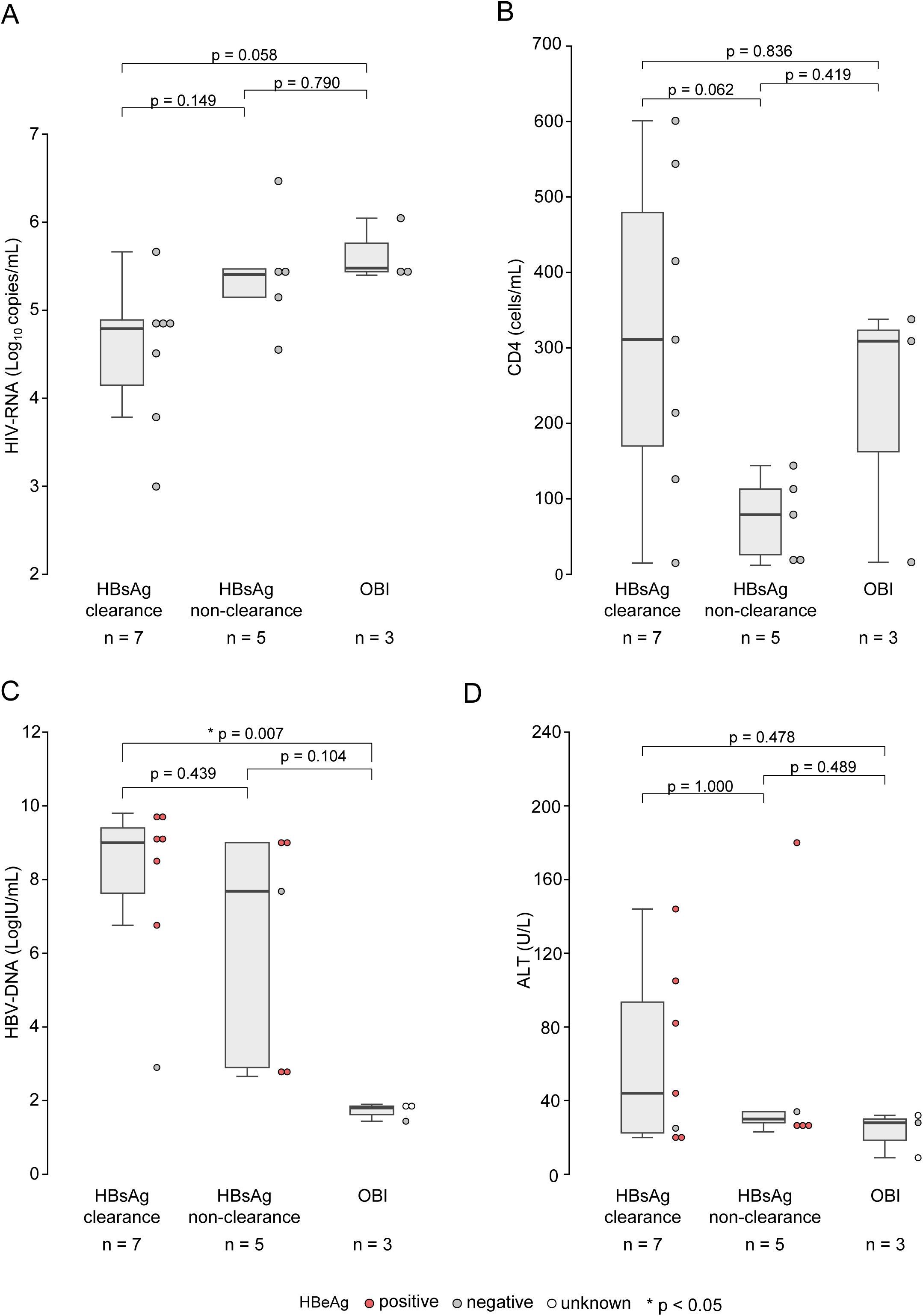
Comparison of (A) HIV-RNA, (B) CD4 count, (C) HBV-DNA, and (D) ALT by HBsAg clearance status (HBsAg clearance, HBsAg non-clearance, and OBI). HBeAg status of individual patients is indicated by dot color. HBV-DNA differed significantly between the HBsAg clearance and OBI groups (\**p* = 0.007); all other pairwise comparisons were not significant.

We successfully sequenced the HBsAg-coding region in all 3 OBI cases (OBI-53, OBI-152, and OBI-168). All three sequences were consistent with genotype C2, the genotype most prevalent in Japan, and were identical to the C2 reference (AB050018) across the sequenced region. However, OBI-152 additionally yielded a second, distinct sequence corresponding to genotype A2, indicating coinfection with two HBV genotypes. Relative to a genotype A2 reference (AB116080), this second sequence carried two amino acid substitutions: Ser45Ala (S45A), outside the MHR, and Asn131Lys (N131K) within the HBsAg ‘a’ determinant (**Figure 4A, E**). Among 4,372 genotype A sequences in HBVdb, asparagine at position 131 was the dominant residue (98.63%), whereas lysine (the OBI-152 variant) was present in only 10 sequences (0.23%) (**Figure 4B**, **C**), indicating that N131K is a rare variant. **Figure 4D** illustrates the three-dimensional location of residue 131 within the ‘a’ determinant loop of the HBsAg monomer, alongside other positions previously reported to influence anti-HBs antibody binding, including residues 123, 126, 129, and 141–145 [14–16].

**Figure 4.**
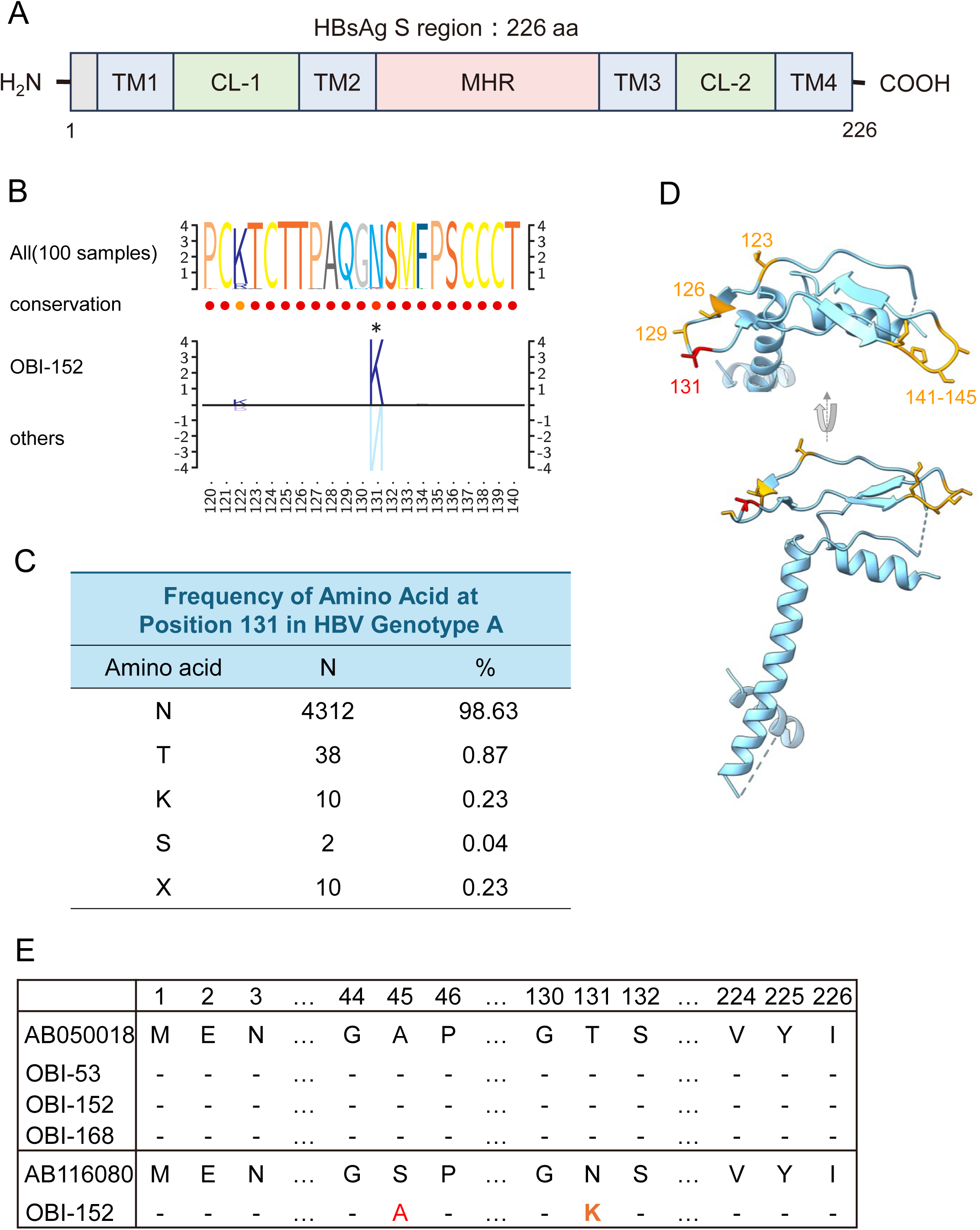
Genetic characterization of the HBsAg S region. (A) Domain structure of S-HBsAg (226 aa) showing transmembrane domains (TM1–TM4), cytoplasmic loops (CL-1, CL-2), and the major hydrophilic region (MHR). (B) Sequence conservation logos for the ‘a’ determinant region (residues 120–140): all 100 reference sequences (top), OBI-152 genotype A2 sequence (middle), and other OBI cases (bottom). Asterisk denotes position 131. (C) Frequency of amino acid residues at position 131 among genotype A HBV sequences in HBVdb. X, ambiguous or undetermined residue. (D) Cartoon representation of the HBsAg monomer, highlighting ‘a’ determinant residues previously reported to affect anti-HBs antibody binding in orange (positions 123, 126, 129, and 141–145) and the site of N131K (residue 131) in red. (E) Amino acid alignment of OBI cases against genotype C2 (AB050018) and genotype A2 (AB116080) references; red letters indicate substitutions unique to OBI-152 (S45A and N131K).

In the loop-completed structural model, the minimum atom-atom distance between residue 131 and the nearest Fab atom was 18.9 Å, indicating that position 131 does not constitute a direct paratope contact (**Figure 5A**). Molecular dynamics simulations revealed a progressive reduction in predicted MM-GBSA binding free energy across constructs: T131 (−56.8 ± 4.4 kcal/mol), N131 (−54.1 ± 3.3 kcal/mol), and K131 (−48.4 ± 2.2 kcal/mol; mean ± SD across independent replicates) (**Figure 5B**). The difference between T131 and K131 was not statistically significant (*p* = 0.069, Welch’s t-test on independent replicates, two-tailed). These findings are consistent with an indirect mechanism in which N131K may alter local loop dynamics rather than directly disrupting the antibody-binding epitope.

**Figure 5.**
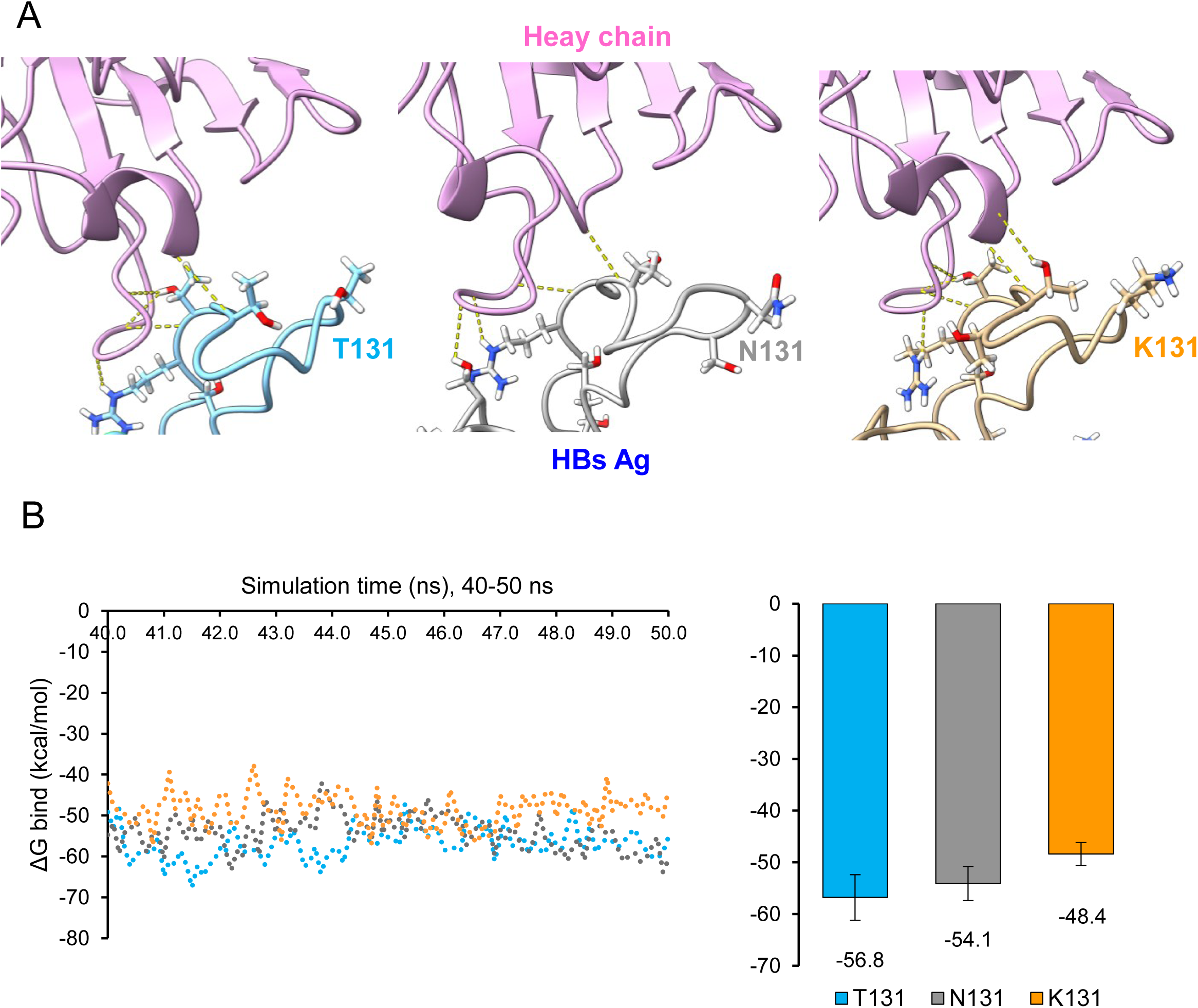
*In silic*o structural and binding-energy analysis of residue 131 at the HBsAg–Fab interface. (A) Loop-completed structural models derived from PDB 9U9B, with residue 131 shown as sticks: T131 (cyan), N131 (gray), and K131 (gold), overlaid on the HBC34 Fab heavy chain (pink). Hydrogen bonds are indicated by dashed yellow lines. The monomer depicted is that in which residue 131 is structurally defined; residues 127–132 are unresolved in the partner monomer in all three Fab-complex structures (9U9B, 9IYY, 9JT1). Residue 131 lies 18.9 Å from the nearest Fab atom and does not constitute a direct paratope contact. (B) MM-GBSA ΔG_bind_ from the last 10 ns of each 50 ns simulation. Left: per-frame time series. Right: mean ΔG_bind_ ± SD across independent replicates (T131, n = 3; N131, n = 3; K131, n = 2). MM-GBSA values without entropy correction may overestimate absolute binding affinity; the relative order across constructs is the meaningful comparison.

## Discussion

To our knowledge, this is the first report to combine clinical factor analysis of HBsAg clearance with genetic and structural characterization of OBI in a Japanese HIV/HBV-coinfected cohort. In this single-center cohort of people with HIV/HBV-coinfection, more than half of HBsAg-positive individuals cleared HBsAg during follow-up, and clearance was concentrated within the first 3–4 years after ART initiation. This temporal pattern, together with the association between clearance and preserved CD4 count and suppressed HIV-RNA, is consistent with a model in which early immune reconstitution (rather than cumulative NRTI exposure duration) drives HBsAg clearance. A recent multisite Asian HIV/HBV cohort study reported HBsAg loss in 22% of patients within 2 years of starting HBV-active ART, with most events occurring by 12 months [17], a kinetic pattern broadly consistent with the early, front-loaded clearance observed here, even though our overall clearance rate (58%) was considerably higher, reflecting our small sample size and single-center cohort. If confirmed in larger cohorts, this early-window pattern would suggest that patients who remain HBsAg-positive beyond the first 3–4 years of ART are unlikely to clear subsequently, informing the timing of any switch to NRTI-sparing regimens.

Baseline HBV-DNA and ALT levels did not differ significantly between the clearance and non-clearance groups. Both HBV-DNA and ALT were numerically higher in the clearance group, and HBV-DNA was significantly higher in the clearance group than in the OBI group. A prior HIV/HBV cohort study also associated HBsAg clearance with higher ALT levels [17]. These findings are consistent with the hypothesis that higher viral replicative activity and higher ALT levels at treatment initiation may be associated with subsequent HBsAg clearance. Our study did not directly measure immune pressure or other determinants of hepatic inflammation and included only a small number of patients, so the underlying mechanism remains uncertain.

Because OBI was identified before ART initiation, the N131K substitution detected in one OBI case may indicate a mechanism distinct from NRTI-mediated suppression of HBV replication. Of the two substitutions identified in OBI-152, N131K is the more likely functional candidate, as it lies within the ‘a’ determinant targeted by most HBsAg immunoassays, and may contribute to false-negative results through altered antigen recognition; S45A, by contrast, lies outside the MHR. Well-characterized substitutions elsewhere in this epitope, such as G145R, are known to abolish binding of multiple monoclonal antibodies and to cause vaccine and immunoprophylaxis escape [15,16], illustrating that single residues within the ‘a’ determinant can have outsized antigenic consequences. To our knowledge, no previously published structural or antibody-binding data exist for a substitution specifically at position 131, in either its wild-type (genotype A2, asparagine) or variant (lysine) form; all cryo-EM structures of HBsAg–antibody complexes reported to date (PDB 9U9B, 9IYY, 9JT1) [14,18] were solved using genotype D-background constructs carrying threonine at this position. This underscores that OBI in HIV-positive patients may be mechanistically heterogeneous, encompassing both assay-detectable viral suppression and genuine immunoassay escape, with potentially different implications for reactivation risk when NRTIs are withdrawn. Consistent with an indirect mechanism, molecular dynamics simulation predicted progressive weakening of binding affinity from T131 through N131 to K131 despite the absence of direct contact between residue 131 and the Fab paratope, suggesting that N131K may perturb local antigenic loop dynamics rather than disrupting a defined epitope contact (**Figure 5B**).

Several limitations should be noted. First, this is a single-center, retrospective study with a small number of patients (12 HBsAg-positive, 3 OBI), limiting statistical power and precluding multivariable analysis; the associations identified should be regarded as hypothesis-generating. Second, because the retrospective design could not fully exclude undiagnosed acute HBV infection at baseline, the true HBsAg clearance rate may have been overestimated. Third, genetic analysis was restricted to the HBsAg-coding region; whole-genome sequencing would allow assessment of mutations elsewhere in the genome, including the basal core promoter and precore regions, that may also contribute to OBI. Fourth, the *in silico* structural analysis is based on multiple independent simulation replicates (T131 and N131, n=3 each; K131, n=2); however, sample sizes remain small and findings should be regarded as hypothesis-generating and require confirmation by experimental binding assays such as surface plasmon resonance or enzyme-linked immunosorbent assay with defined monoclonal antibodies. Finally, this study did not assess HBV reactivation after NRTI withdrawal; prospective evaluation of reactivation risk stratified by HBsAg clearance status and genetic findings is an important next step.

In conclusion, HBsAg clearance was common among HIV/HBV-coinfected patients in this cohort and was associated with preserved baseline immune status and higher baseline HBV replicative activity, with most clearance events occurring within the first 3–4 years of ART. A rare substitution (N131K) in the HBsAg antigenic loop was identified as a plausible contributor to in one OBI case, within a genotype A2 sequence that also carried a second substitution (S45A) outside the principal antibody-binding region. These findings may inform both the timing of NRTI-sparing regimen switches and the need for HBV-DNA testing in HBsAg-negative patients prior to such transitions.

## Data Availability

All data produced in the present work are contained in the manuscript

## Acknowledgements

We are grateful to all patients followed at the HIV center of Kumamoto University Hospital for their cooperation and the use of their clinical data. We thank the nursing and clinical staff of the Department of Hematology, Rheumatology, and Infectious Diseases for their dedicated care of the cohort.

## Funding

This research did not receive any specific grant from funding agencies in the public, commercial, or not-for-profit sectors.

## Ethical approval

The study protocol was approved by the Ethics Committee of Kumamoto University (Approval No. 1942).

## Declaration of competing interest

The authors have no relevant financial or non-financial interests to disclose.

## Author contributions

Conceptualization: MT, TN; Methodology: MT, TN; Software: MT, TN; Formal analysis and investigation: MT, TN; Resources: HN, SM; Writing – original draft: MT, TN; Writing – review and editing: HN, JY, SM; Visualization: MT, TN; Supervision: SM. All authors read and approved the final manuscript.

## Declaration of generative AI use

During the preparation of this work, the authors used Claude Sonnet 5 to correct spelling and grammatical errors, improve English readability, and assist with data organization and figure preparation. After using this tool/service, the authors reviewed and edited the content as needed and take full responsibility for the content of the published article.

## Notes

### Competing Interest Statement

The authors have declared no competing interest.

### Author Declarations

The study protocol was approved by the Ethics Committee of Kumamoto University (Approval No. 1942).

